# Diet quality and risk and severity of COVID-19: a prospective cohort study

**DOI:** 10.1101/2021.06.24.21259283

**Authors:** Jordi Merino, Amit D. Joshi, Long H. Nguyen, Emily R. Leeming, Mohsen Mazidi, David A Drew, Rachel Gibson, Mark S. Graham, Chun-Han Lo, Joan Capdevila, Benjamin Murray, Christina Hu, Somesh Selvachandran, Sohee Kwon, Wenjie Ma, Cristina Menni, Alexander Hammers, Shilpa N. Bhupathiraju, Shreela V. Sharma, Carole Sudre, Christina M. Astley, Walter C. Willet, Jorge E. Chavarro, Sebastien Ourselin, Claire J. Steves, Jonathan Wolf, Paul W. Franks, Tim D. Spector, Sarah E. Berry, Andrew T. Chan

## Abstract

**Objective:** Poor metabolic health and certain lifestyle factors have been associated with risk and severity of coronavirus disease 2019 (COVID-19), but data for diet are lacking. We aimed to investigate the association of diet quality with risk and severity of COVID-19 and its intersection with socioeconomic deprivation.

**Design:** We used data from 592,571 participants of the smartphone-based COVID Symptom Study. Diet quality was assessed using a healthful plant-based diet score, which emphasizes healthy plant foods such as fruits or vegetables. Multivariable Cox models were fitted to calculate hazard ratios (HR) and 95% confidence intervals (95% CI) for COVID-19 risk and severity defined using a validated symptom-based algorithm or hospitalization with oxygen support, respectively.

**Results:** Over 3,886,274 person-months of follow-up, 31,815 COVID-19 cases were documented. Compared with individuals in the lowest quartile of the diet score, high diet quality was associated with lower risk of COVID-19 (HR, 0.91; 95% CI, 0.88-0.94) and severe COVID-19 (HR, 0.59; 95% CI, 0.47-0.74). The joint association of low diet quality and increased deprivation on COVID-19 risk was higher than the sum of the risk associated with each factor alone (*P*_*interaction*_=0.005). The corresponding absolute excess rate for lowest vs highest quartile of diet score was 22.5 (95% CI, 18.8-26.3) and 40.8 (95% CI, 31.7-49.8; 10,000 person-months) among persons living in areas with low and high deprivation, respectively.

**Conclusions:** A dietary pattern characterized by healthy plant-based foods was associated with lower risk and severity of COVID-19. These association may be particularly evident among individuals living in areas with higher socioeconomic deprivation.

## Introduction

Poor metabolic health^1,2^ has been associated with increased risk and severity of coronavirus disease 2019 (COVID-19), and excess adiposity or preexisting liver disease might be causally associated with increased risk of death from COVID-19.^3,4^ Underlying these conditions is the contribution of a diet, which may be independently associated with COVID-19 risk and severity.

On the basis of prior scientific evidence, diet quality scores have been developed to evaluate the healthfulness of dietary patterns.^5–7^ Dietary patterns capture the complexity of food intakes better than any one individual food item and offer the advantage of describing usual consumption of foods in typical diets.^8^ One such diet score is the healthful plant-based diet index (hPDI), which emphasizes intake of healthy plant foods, such as fruits, vegetables, and whole grains, and has been associated with lower risk of metabolic diseases.^5,9,10^

Adherence to healthful dietary patterns may also be a proximal manifestation of distal social determinants of health.^11–13^ Addressing adverse social determinants of health, such as poor nutrition, has been shown to reduce the burden of certain infectious diseases in the past,^14^ supporting calls for prioritizing social determinants of health in the public health response to COVID-19. However, evidence on the association between diet quality and the risk and severity of COVID-19 is lacking, especially in the context of upstream social determinants of health. To address this evidence gap, we analyzed data for 592,571 United Kingdom (UK) and United States (US) participants from the smartphone-based COVID Symptom Study,^15^ to prospectively investigate the association of diet quality with risk and severity of COVID-19 and its intersection with socioeconomic deprivation.

## Materials and Methods

### Study design and participants

The COVID Symptom Study is a smartphone-based study conducted in the UK and US. Study design and sampling procedures have been published elsewhere.^15^ This analysis included participants recruited from March 24, 2020 and followed until December 2, 2020. Participants who reported any symptoms related to COVID-19 prior to start of follow-up, or reported symptoms that classified them as having predicted COVID-19 within 24 hours of first entry, or who tested positive for COVID-19 at any time prior to start of follow-up or 24 hours after first entry were excluded. We also excluded participants younger than 18 years old, pregnant, and participants who logged only one daily assessment during follow-up. At enrollment, we obtained informed consent to the use of volunteered information for research purposes and shared relevant privacy policies and terms of use agreements. The study protocol was approved by the Mass General Brigham Human Research Committee (protocol 2020P000909) and King’s College London Ethics Committee (REMAS ID 18210, LRS-19/20-18210).

### Data collection procedures

Information on demographic factors was collected through standardized questionnaires at baseline,^15^ including self-reported COVID-19 or any COVID-19 related symptoms and personal medical history including lung disease, diabetes, cardiovascular disease, cancer, kidney disease, and use of medications. During follow-up, daily prompts queried for updates on interim symptoms, health care visits, and COVID-19 testing results. Through software updates, a survey to examine self-reported diet and lifestyle habits during the pre/early-pandemic period was launched between August and September 2020. Details about this survey are available in the Supplementary Methods and published elsewhere.^16^

### Assessment of diet quality

Diet quality was assessed using information obtained from an amended version of the Leeds Short Form Food Frequency Questionnaire^17^ that included 27 food items (online supplementary methods). Participants were asked how often on average they had consumed one portion of each item in a typical week. The responses had eight frequency categories ranging from “rarely or never” to “five or more times per day”.

Diet quality was quantified using the validated hPDI score.^5^ To compute the hPDI, the 27 food items were combined into 14 food groups (online supplementary table 1). The original hPDI score included 18 food groups but nuts, vegetable oils, tea or coffee, and animal fat were not specifically queried. Food groups were ranked into quintiles and given positive (healthy plant food groups) or reverse scores (less healthy plant and animal food groups). With positive scores, participants within the highest quintile of a food group received a score of 5, following on through to participants within the lowest quintile who received a score of 1. With reverse scores, this pattern of scoring was inverted. All component scores were summed to obtain a total score ranging from 14 (lowest diet quality) to 70 (highest) points. Criteria for generation of the hPDI are provided in online supplemental table 2. As an additional method to quantify diet quality based on available diet information, we used the Diet Quality Score (DQS).^17^ The DQS is a score for adherence to UK dietary guidelines and was computed from five broad categories including fruits, vegetables, total fat, oily fish, and non-milk extrinsic sugars. Each component was scored from 1 (unhealthiest) to 3 (healthiest) points, with intermediate values scored proportionally (online supplementary table 3). All component scores were summed to obtain a total score ranging from 5 (lowest diet quality) to 15 (highest) points (online supplementary table 4).

### Assessment of COVID-19 risk and severity

The primary outcome of this analysis was COVID-19 risk defined using a validated symptom-based algorithm,^18^ which provides similar estimates of COVID-19 prevalence and incidence as those reported from the Office for National Statistics Community Infection Survey.^19^ Details on the symptoms included in the predictive algorithm and corresponding weights are provided in the online supplementary methods. In brief, the symptom-based approach uses an algorithm to predict whether a participant has been infected with SARS-CoV-2 on the basis of their reported symptoms, age, and sex. The rationale for symptom-based classifier as a primary outcome was due to widespread difficulties obtaining testing during the early stages of the pandemic.^20^ Secondary outcomes were confirmed COVID-19 based on a self-report of a reverse transcription polymerase chain reaction (RT-PCR) positive test and COVID-19 severity. COVID-19 severity was ascertained based on a report of the need for a hospital visit which required 1) non-invasive breathing support, 2) invasive breathing support, and 3) administration of antibiotics combined with oxygen support (online supplementary methods).

### Statistical analysis

We summarized continuous measurements by using medians and interquartile ranges, and present categorical observations as frequency and percentages. Based on zip code (US) or post code (UK) of residence, participants were assigned to country-specific community-level socioeconomic measures including socioeconomic deprivation and population density (online supplementary methods). The methods for classifying socioeconomic deprivation, population density, and other *a priori* selected covariates are provided in online supplementary methods. Multiple imputations by chained equations with five imputations were used to impute missing values. All covariates in the primary analysis were included in the multiple imputation procedure, and estimates generated from each imputed dataset were combined using Rubin’s rules.^21^

Follow-up time for each participant started 24 hours after first log-in to the time of predicted COVID-19 (or to time of secondary outcomes) or date of last entry prior to December 2, 2020, whichever occurred first. We modeled the diet quality score as a continuous variable and generated categories of the score based on quartiles of the distribution (quartile 1, low diet quality; quartiles 2-3, intermediate diet quality; quartile 4, high diet quality). Cox regression models stratified by calendar date at study entry, country of origin, and 10-year age group were used to calculate hazard ratio (HR) and 95% confidence intervals (95% CI) for COVID-19 risk and severity (age-adjusted model 1). Model 2 was further adjusted for sex, race/ethnicity, index of multiple deprivation, population density, presence of diabetes, cardiovascular disease, lung disease, cancer, kidney disease, and healthcare worker status. Model 3 was further adjusted for body mass index, smoking status, and physical activity. We verified the proportional hazards assumption of the Cox model by using the Schoenfeld residuals technique.^22^ Absolute risk was calculated as the percentage of COVID-19 cases occurring per 10,000 person-months in a given group. We used restricted cubic splines with four knots (at the 2.5th, 25th, 75th, and 97.5th percentiles) to assess for non-linear associations between diet quality and COVID-19 risk.

In secondary analyses, we used a self-report of a positive test to define COVID-19 risk. For these analyses, we used inverse probability-weighted Cox models to account for predictors of obtaining country-specific testing. Inverse probability-weighted analyses included presence of COVID-19-related symptoms, interaction with a person with COVID-19, occupation as a healthcare worker, age group, and race. Inverse probability-weighted Cox models were stratified by 10-year age group and date with additional adjustment for the covariates used in previous models. For severe COVID-19 analyses, we adjusted for the same covariates used in previous models. As an additional method to quantify diet quality we used the DQS and tested for associations between diet quality and COVID-19 risk and severity. In addition, we censored our analyses to cases that occurred after completing the diet survey to investigate potential bias due to time-varying confounding.

In subgroup analyses, we assessed the association between diet quality and COVID-19 risk according to comorbidities, demographic, and lifestyle characteristics. We also classified participants according to categories of the diet quality score and socioeconomic deprivation (nine categories based on thirds of diet quality score and deprivation index) and conducted joint analyses for COVID-19 risk. We tested for additive interactions by assessing the relative excess risk due to interaction, and further examined the COVID-19 risk proportions attributable to diet, deprivation, and to their interaction (online supplementary methods).^23^

We conducted sensitivity analyses to account for regional differences in the effective reproductive number (*R*_*t*_) or other risk mitigating behaviors such as mask wearing. Details on how we obtained and classified individuals for these analyses are provided in online supplementary methods. Two-sided p-values of <0.05 were considered statistically significant for main analyses. All statistical analyses were performed using R software, version 4.0.3 (R Foundation).

## Results

Self-reported diet quality was evaluated in 647,137 survey responders, of which 54,566 were excluded due to prevalent COVID (n=1,555), presence of any symptoms at baseline (n=47,594), logged only once (n=1,201), pregnancy (n=1,129), or age under 18 year (n=3,087; online supplementary figure 1). Baseline characteristics of the 592,571 participants included in this study according to categories of the hPDI score are shown in table 1. Participants in the highest quartile of the diet score (reflecting a healthier diet) were more likely than participants in the lowest quartile to be older, female, healthcare workers, of lower BMI, engage in physical activities ≥ 5 days/week, and less likely to reside in areas with higher socioeconomic deprivation. The hPDI score was normally distributed (online supplementary figure 2).

**Table 1.**
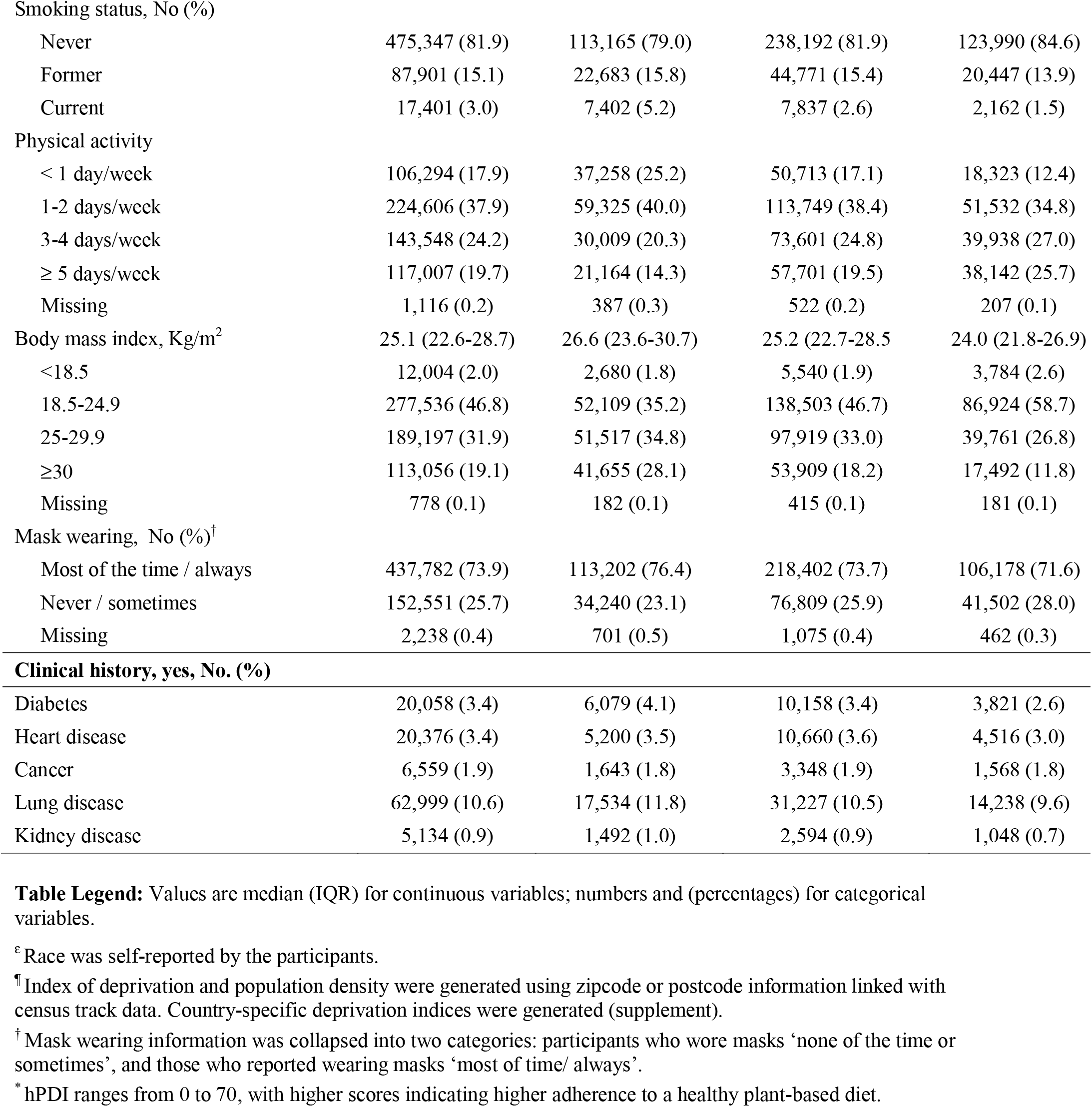
Baseline characteristics of study participants according to categories of the diet quality score

|  | All participants<br>(n=592,571) | Low hPDI<br>(Q1; n=148,143) | Intermediate hPDI<br>(Q2-Q3; n=296,286) | High hPDI<br>(Q4; n=148,142) |
| --- | --- | --- | --- | --- |
| hPDI score, median [IQR] | 50 (47 to 54) | 45 (43-47) | 51 (49-52) | 56 (55-58) |
| **Demographic characteristics** |  |  |  |  |
| Age, years | 56 (44-65) | 52 (41-62) | 57 (45-66) | 57 (45-65) |
| ≥18-24 | 14,397 (2.4) | 5,146 (3.4) | 5,846 (2.0) | 3,405 (2.3) |
| 25-34 | 52,922 (8.9) | 16,535 (11.2) | 23,150 (7.8) | 13,237 (8.9) |
| 35-44 | 86,251 (14.6) | 26,907 (18.2) | 40,145 (13.5) | 19,199 (13.0) |
| 45-54 | 125,802 (21.2) | 34,890 (23.6) | 62,491 (21.1) | 28,421 (19.2) |
| 55-64 | 158,637 (26.8) | 34,279 (23.1) | 81,837 (27.6) | 42,521 (28.7) |
| ≥65 | 153,810 (26.0) | 30,215 (20.4) | 82,413 (27.8) | 41,182 (27.8) |
| Missing | 752 (0.1) | 171 (0.1) | 404 (0.1) | 177 (0.1) |
| Sex, No. (%) |  |  |  |  |
| Male | 187,450 (31.6) | 58,199 (39.3) | 93,162 (31.4) | 36,089 (24.4) |
| Female | 404,126 (68.2) | 89,706 (60.5) | 202,605 (68.4) | 111,815 (75.5) |
| Prefer not to say | 995 (0.2) | 238 (0.2) | 519 (0.2) | 238 (0.2) |
| Race<sup>e</sup>, No. (%), |  |  |  |  |
| White | 568,770 (96.0) | 141,365 (95.4) | 284,804 (96.1) | 142,601 (96.3) |
| Black | 4,328 (0.7) | 1,466 (1.0) | 2,053 (0.7) | 809 (0.5) |
| Asian | 10,435 (1.8) | 2,954 (1.9) | 5,043 (1.7) | 2,438 (1.6) |
| Other | 7,228 (1.2) | 1,925 (1.3) | 3,463 (1.2) | 1,840 (1.2) |
| Missing | 1,810 (0.3) | 433 (0.3) | 923 (0.3) | 454 (0.3) |
| Country, No. (%) |  |  |  |  |
| UK | 543,984 (91.8) | 135,360 (91.4) | 272,494 (92.0) | 136,130 (91.9) |
| US | 48,587 (8.2) | 12,783 (8.6) | 23,792 (8.0) | 12,012 (8.1) |
| Index of deprivation, No. (%)<sup>¶</sup> |  |  |  |  |
| Most deprived, decile 1 | 1,3416 (2.3) | 4,696 (3.1) | 6,163 (2.1) | 2,557 (1.7) |
| Least deprived, decile 10 | 103,608 (17.5) | 23,122 (15.6) | 53,652 (18.1) | 26,834 (18.1) |
| Missing | 40,759 (6.9) | 10,489 (7.1) | 20,249 (6.8) | 10,021 (6.8) |
| Population density, km<sup>2</sup>, No. (%)<sup>¶</sup> |  |  |  |  |
| <500 | 119,782 (20.2) | 28,139 (19.0) | 61,230 (20.7) | 30,413 (20.5) |
| 500-1,999 | 90,541 (15.3) | 23,631 (16.0) | 45,902 (15.5) | 21,008 (14.2) |
| 2,000 4,999 | 94,345 (15.9) | 24,813 (16.7) | 47,233 (15.9) | 22,299 (15.1) |
| ≥5,000 | 244,295 (41.2) | 60,156 (40.6) | 120,319 (40.6) | 63,820 (43.1) |
| Missing | 43,608 (7.4) | 11,404 (7.7) | 21,602 (7.3) | 10,602 (7.2) |
| Healthcare worker, yes, No. (%) | 41,141 (6.9) | 10,633 (2.3) | 20,183 (6.8) | 10,325 (7.0) |
| **Lifestyle characteristics** |  |  |  |  |
| Smoking status, No (%) |  |  |  |  |
| Never | 475,347 (81.9) | 113,165 (79.0) | 238,192 (81.9) | 123,990 (84.6) |
| Former | 87,901 (15.1) | 22,683 (15.8) | 44,771 (15.4) | 20,447 (13.9) |
| Current | 17,401 (3.0) | 7,402 (5.2) | 7,837 (2.6) | 2,162 (1.5) |
| Physical activity |  |  |  |  |
| < 1 day/week | 106,294 (17.9) | 37,258 (25.2) | 50,713 (17.1) | 18,323 (12.4) |
| 1-2 days/week | 224,606 (37.9) | 59,325 (40.0) | 113,749 (38.4) | 51,532 (34.8) |
| 3-4 days/week | 143,548 (24.2) | 30,009 (20.3) | 73,601 (24.8) | 39,938 (27.0) |
| ≥ 5 days/week | 117,007 (19.7) | 21,164 (14.3) | 57,701 (19.5) | 38,142 (25.7) |
| Missing | 1,116 (0.2) | 387 (0.3) | 522 (0.2) | 207 (0.1) |
| Body mass index, Kg/m<sup>2</sup> |  |  |  |  |
| <18.5 | 12,004 (2.0) | 2,680 (1.8) | 5,540 (1.9) | 3,784 (2.6) |
| 18.5-24.9 | 277,536 (46.8) | 52,109 (35.2) | 138,503 (46.7) | 86,924 (58.7) |
| 25-29.9 | 189,197 (31.9) | 51,517 (34.8) | 97,919 (33.0) | 39,761 (26.8) |
| ≥30 | 113,056 (19.1) | 41,655 (28.1) | 53,909 (18.2) | 17,492 (11.8) |
| Missing | 778 (0.1) | 182 (0.1) | 415 (0.1) | 181 (0.1) |
| Mask wearing, No (%)<sup>†</sup> |  |  |  |  |
| Most of the time / always | 437,782 (73.9) | 113,202 (76.4) | 218,402 (73.7) | 106,178 (71.6) |
| Never / sometimes | 152,551 (25.7) | 34,240 (23.1) | 76,809 (25.9) | 41,502 (28.0) |
| Missing | 2,238 (0.4) | 701 (0.5) | 1,075 (0.4) | 462 (0.3) |
| **Clinical history, yes, No. (%)** |  |  |  |  |
| Diabetes | 20,058 (3.4) | 6,079 (4.1) | 10,158 (3.4) | 3,821 (2.6) |
| Heart disease | 20,376 (3.4) | 5,200 (3.5) | 10,660 (3.6) | 4,516 (3.0) |
| Cancer | 6,559 (1.9) | 1,643 (1.8) | 3,348 (1.9) | 1,568 (1.8) |
| Lung disease | 62,999 (10.6) | 17,534 (11.8) | 31,227 (10.5) | 14,238 (9.6) |
| Kidney disease | 5,134 (0.9) | 1,492 (1.0) | 2,594 (0.9) | 1,048 (0.7) |

**Table Legend:** Values are median (IQR) for continuous variables; numbers and (percentages) for categorical variables.
<sup>ε</sup> Race was self-reported by the participants.
<sup>¶</sup> Index of deprivation and population density were generated using zipcode or postcode information linked with census track data. Country-specific deprivation indices were generated (supplement).
<sup>†</sup> Mask wearing information was collapsed into two categories: participants who wore masks ‘none of the time or sometimes’, and those who reported wearing masks ‘most of time/ always’.
<sup>\*</sup> hPDI ranges from 0 to 70, with higher scores indicating higher adherence to a healthy plant-based diet.

Over 3,886,274 person-months of follow-up, 31,815 COVID-19 cases were documented. Crude COVID-19 rates per 10,000 person-months were 72.0 (95% CI, 70.4-73.7) for participants in the highest quartile of the diet score and 104.1 (95% CI, 101.9-106.2) for those in the lowest quartile. The corresponding age-adjusted HR for COVID-19 risk was 0.80 (95% CI, 0.78-0.83, table 2). Differences in the risk of COVID-19 persisted after adjustment for potential confounders. In fully adjusted models, the multivariable-adjusted HR for COVID-19 risk was 0.91 (95% CI, 0.88-0.94) when we compared participants with high diet quality to those with low diet quality. We observed non-linear decreasing trends in the risk of COVID-19 with higher diet quality (*P* < 0.001 for non-linearity), in which COVID-19 risk plateau among individuals with a diet quality score > 50 (online supplementary figure 3). The association between diet quality and COVID-19 risk was consistent but attenuated in secondary analyses using the DQS score (HR, 0.92; 95% CI, 0.89-0.95; online supplementary table 5), and became non-significant in fully adjusted models (HR, 1.00; 95% CI, 0.97-1.03). We also investigated whether our primary findings were consistent in an analysis censored to cases that occurred after the completion of the diet survey. These analyses showed that high diet quality, compared to low diet quality, was associated with lower COVID-19 risk (multivariable-adjusted HR 0.88; 95% CI, 0.83-0.93; online supplementary table 6).

**Table 2.** Adjusted hazard ratios of COVID-19 risk and severity according to healthful plant-based dietary index scores.

|  | **Low hPDI**<br>(Q1; n=148,143) | **Intermediate hPDI**<br>(Q2-Q3; n=296,286) | **High hPDI**<br>(Q4; n=148,142) | **P for trend** |
| --- | --- | --- | --- | --- |
| hPDI score, median (IQR) | 45 (43-47) | 51 (49-52) | 56 (55-58) |  |
| **COVID-19 risk** |  |  |  |  |
| No. of events/person-months | 8,739 / 839,747 | 15,733 / 2,026,824 | 7,359 / 1,022,078 | — |
| Incidence rate (10,000 person-months; 95% CI) | 104.1 (101.9-106.2) | 77.6 (76.4-78.8) | 72.0 (70.4-73.7) | — |
| Age-adjusted model | 1.00 (Ref) | 0.85 (0.82-0.87) | 0.80 (0.78-0.83) | <0.001 |
| Multivariable model 2 | 1.00 (Ref) | 0.85 (0.83-0.87) | 0.81 (0.78-0.83) | <0.001 |
| Multivariable model 3 | 1.00 (Ref) | 0.91 (0.89-0.93) | 0.91 (0.88-0.94) | <0.001 |
| **COVID-19 risk (positive test)** |  |  |  |  |
| No. of events/person-months | 1,423 / 869,664 | 2,829 / 2,081,970 | 1,350 / 1,046,887 | — |
| Incidence rate (10,000 person-months; 95% CI) | 16.4 (15.5-17.2) | 13.6 (13.1-14.1) | 12.9 (12.2-13.6) | — |
| Age-adjusted model<sup>s</sup> | 1.00 (Ref) | 0.86 (0.83-0.90) | 0.79 (0.75-0.83) | <0.001 |
| Multivariable model 2<sup>s</sup> | 1.00 (Ref) | 0.87 (0.84-0.91) | 0.80 (0.76-0.84) | <0.001 |
| Multivariable model 3<sup>s</sup> | 1.00 (Ref) | 0.88 (0.85-0.92) | 0.82 (0.78-0.86) | <0.001 |
| **Severe COVID-19** |  |  |  |  |
| No. of events/person-months | 187 / 871,995 | 390 / 2,086,790 | 163 / 1,049,476 | — |
| Incidence rate (10,000 person-months; 95% CI) | 2.1 (1.9-2.5) | 1.9 (1.7-2.1) | 1.6 (1.3-1.8) | — |
| Age-adjusted model | 1.00 (Ref) | 0.66 (0.56-0.77) | 0.45 (0.36-0.57) | <0.001 |
| Multivariable model 2 | 1.00 (Ref) | 0.66 (0.57-0.78) | 0.45 (0.36-0.57) | <0.001 |
| Multivariable model 3 | 1.00 (Ref) | 0.77 (0.66-0.91) | 0.59 (0.47-0.74) | <0.001 |

**Table Legend:** Hazards ratios and 95% CI for COVID-19 risk and severity. COVID-19 risk defined using a validated symptom-based model. COVID-19 or a RT-PCR positive test report. COVID-19 severity was defined based on hospitalization with requirement of oxygen support (methods, supplement).
Cox proportional hazards models were stratified by calendar date at study entry, country of origin, and 10-year age group (Age-adjusted model).
Multivariable model 2 was further adjusted for sex (male, female), race/ethnicity (White, Black, Asian, Other), index of multiple deprivation (most deprived <3, intermediate deprived 3 to 7, less deprived >7), population density (<500 individuals/km<sup>2</sup>, 500 to 1,999 individuals/km<sup>2</sup>, 2,000 to 4,999 individuals/km<sup>2</sup>, and ≥ 5,000 individuals/km<sup>2</sup>), and healthcare worker status (yes with interaction with COVID-19 patients, yes without interaction with COVID-19 patients, no).
Model 3 was further adjusted for presence of comorbidities [diabetes (yes, no), cardiovascular disease (yes, no), lung disease (yes, no), cancer (yes, no), kidney disease (yes, no)], body mass index (<18.5 kg/m<sup>2</sup>, 18.5 to 24.9 kg/m<sup>2</sup>, 25.0 to 29.9 kg/m<sup>2</sup>, and ≥30 kg/m<sup>2</sup>), smoking status (yes, no), and physical activity (<1 day/week, 1 to 2 days/week, 3 to 4 days/week, ≥5 days/week).
<sup>§</sup> Inverse probability-weighted analyses were conducted to account for predictors of obtaining RT-PCR testing (presence of COVID-19-related symptoms, interaction with a COVID-19 case, healthcare worker, age group, and race). inverse probability-weighted Cox proportional hazards models were stratified by 10-year age group and date with additional adjustment for the covariates used in previous models.

In secondary analyses for COVID-19 risk based on a positive test, we showed that crude COVID-19 incidence rates per 10,000 person-months were 12.9 (95% CI 12.2-13.6) for individuals with high diet quality and 16.4 (95% CI 15.5-17.2) for individuals with low diet quality. The corresponding multivariable-adjusted HR for risk of COVID-19 was 0.82 (95% CI, 0.78-0.86; table 2). For risk of severe COVID-19, crude incidence rates were lower for individuals reporting high diet quality compared to those with low diet quality (1.6 (95% CI, 1.3-1.8) vs. 2.1 (95% CI, 1.9-2.5; per 10,000 person-months) table 2). In the fully adjusted model, high diet quality, as compared to low diet quality, was associated lower risk of severe COVID-19 with an a HR of 0.59 (95% CI, 0.47-0.74; table 2).

In stratified analyses, the inverse association between diet quality and COVID-19 risk was more evident in participants living in areas of high socioeconomic deprivation and those reporting low physical activity levels (*P* < 0.05; table 3). We found no significant effect modification for other characteristics such age, BMI, race/ethnicity or population density. When diet quality and socioeconomic deprivation were combined, there was a risk gradient with low diet quality and high socioeconomic deprivation. Compared with individuals living in areas with low socioeconomic deprivation and high diet quality, the multivariable-adjusted HR for risk of COVID-19 for low diet quality was 1.08 (95% CI, 1.03-1.14) among those living in areas with low socioeconomic deprivation, 1.23 (95% CI, 1.17-1.29) for those living in areas with intermediate socioeconomic deprivation, and 1.47 (95% CI, 1.38-1.52) for those living in areas with high socioeconomic deprivation (figure 1). The joint associations of diet quality and socioeconomic deprivation was higher than the sum of the risk associated with each factor alone (relative excess risk due to interaction (RERI) = 0.05 (95% CI 0.02-0.08); *P*_*interaction*_=0.005; online supplementary table 7). The proportion of contribution to excess COVID-19 risk was estimated to be 31.9% (95% CI, 18.2-45.6) to diet quality, 38.4% (95% CI, 26.5-50.3) to socioeconomic deprivation, and 29.7% (95% CI, 2.1-57.3) to their interaction. The absolute excess rate of COVID-19 per 10,000 person-months for lowest vs highest quartile of the diet score was 22.5 (95% CI, 18.8-26.3) among individuals living in areas with low socioeconomic deprivation and 40.8 (95% CI, 31.7-49.8) among individuals living in areas with high deprivation (online supplementary figure 4)

**Figure 1.**
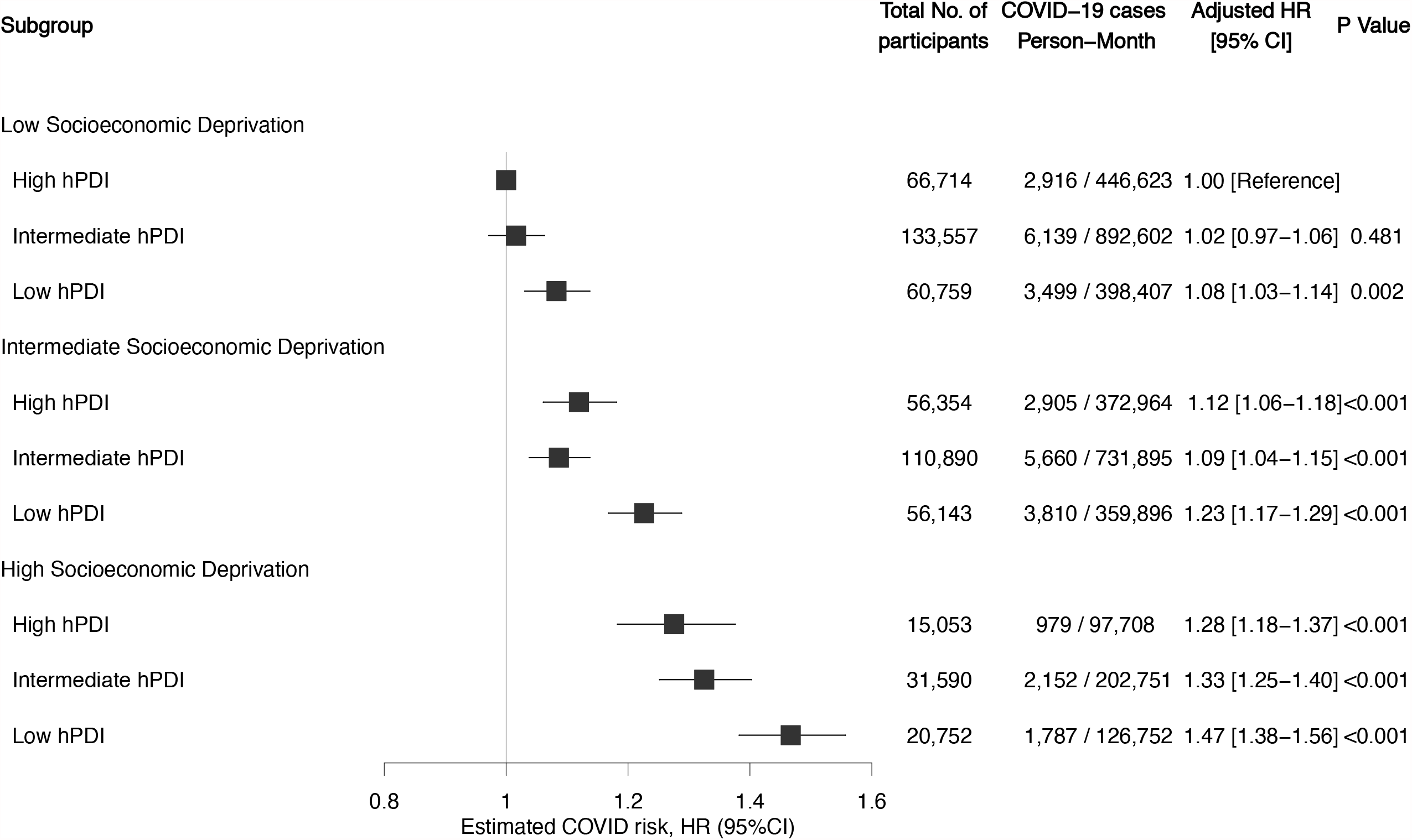
Title: Risk of COVID-19 according to diet quality and socioeconomic deprivation. **Figure legend:** Shown are adjusted hazard ratios and 95% confidence interval of the estimate for predicted COVID-19 according to categories of diet quality and socioeconomic deprivation. Cox model stratified by calendar date at study entry, country of origin, and 10-year age group, and adjusted for sex, race/ethnicity, index of multiple deprivation, population density, presence of diabetes, cardiovascular disease, lung disease, cancer, kidney disease, healthcare worker status, body mass index, smoking status, and physical activity. In these comparisons, participants with high-quality diet and low socioeconomic deprivation served as the reference group.

**Table 3.**
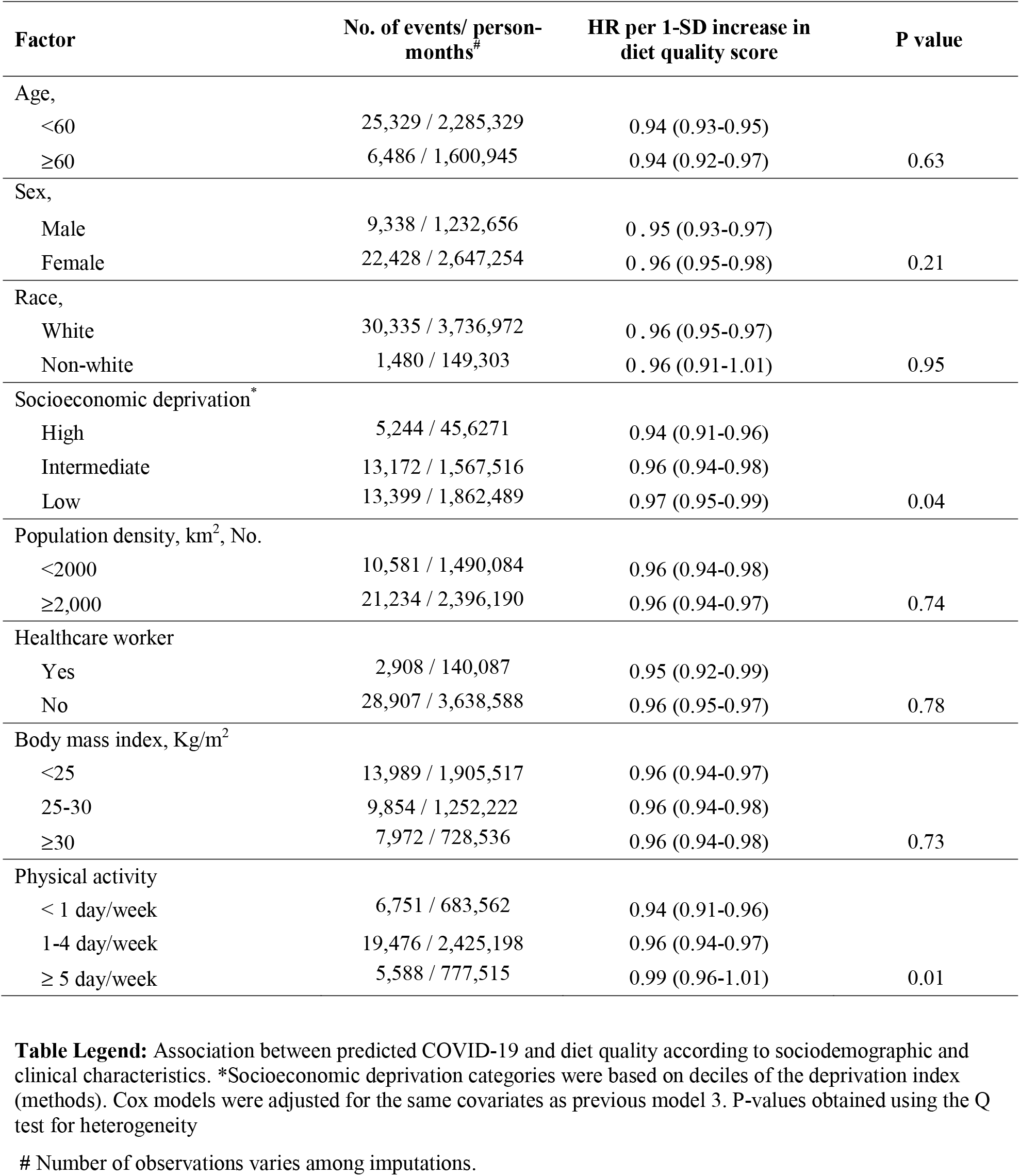
Adjusted hazard ratios of COVID risk according to healthful plant-based dietary index scores stratified by sociodemographic and clinical characteristics.

We conducted a series of sensitivity analyses to further account for variation in *R*_*t*_ or mask wearing. For peak *R*_*t*_ censored analyses, crude COVID-19 rates per 10,000 person-months were 148.1 (95% CI, 139.9-156.8) among participants with low diet quality and 92.9 (95% CI, 86.6-99.5) for participants with high diet quality. The corresponding multivariable-adjusted HR was 0.84 (95% CI, 0.76-0.92, figure 2). The same trend was observed for nadir *R*_*t*_ censored analyses, in which crude COVID-19 rates per 10,000 person-months were 67.1 (95% CI, 61.7-73.0) among participants with low diet quality and 45.8 (95% CI, 41.3-50.5) for participants with high diet quality (multivariable-adjusted HR, 0.89; 95% CI, 0.80-1.00, figure 2). We further adjusted our models for mask wearing. This analysis showed that high diet quality, as compared to low diet quality, was associated with lower risk of COVID-19 with an adjusted HR of 0.88 (95% CI, 0.83-0.94; online supplementary table 8).

**Figure 2.**
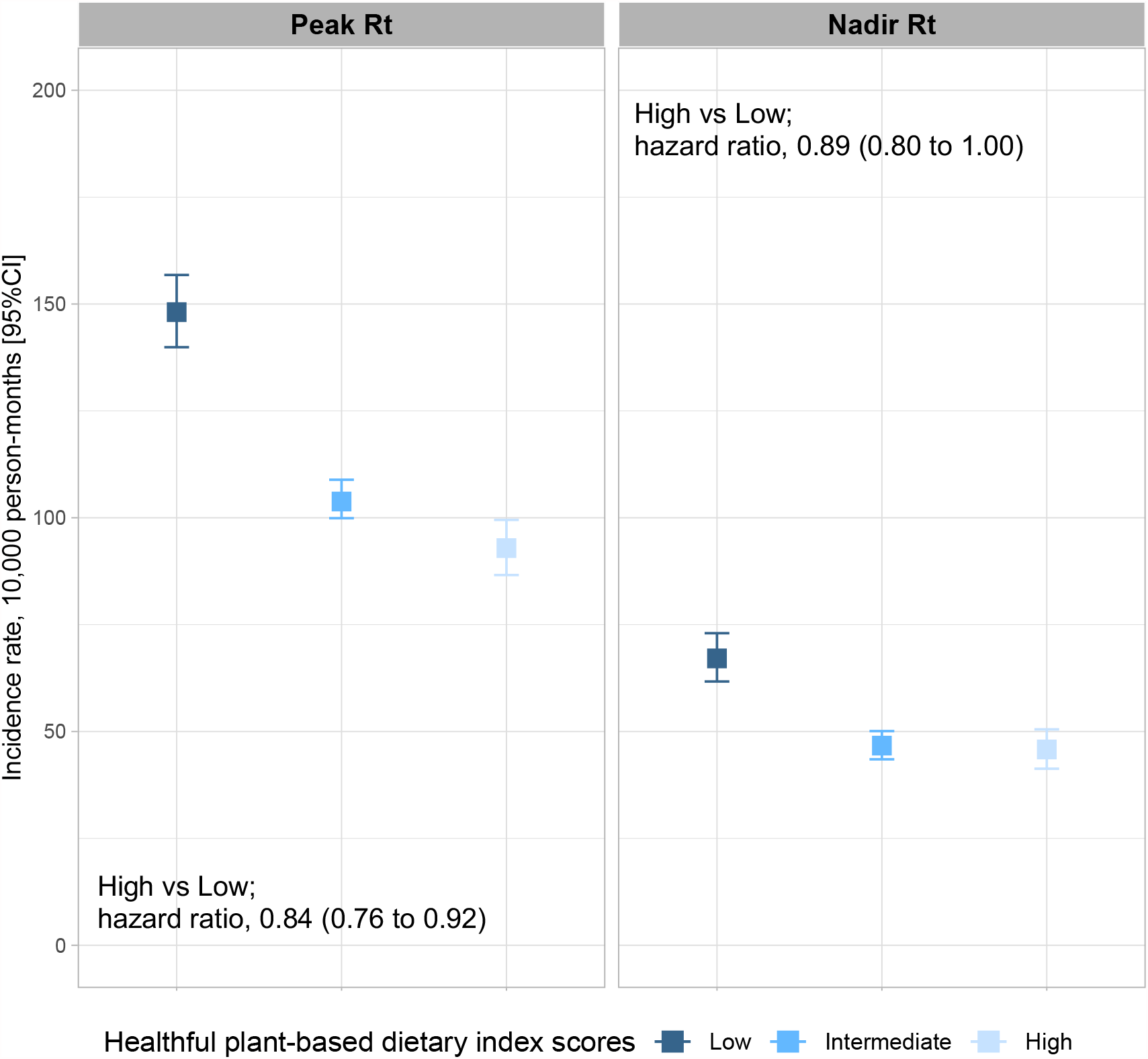
Title: Risk of COVID-19 according to community transmission rate and diet quality. **Figure legend:** COVID-19 incidence rate per 10,000 person-month and 95% confidence interval of the estimate based on different community transmission rate and diet quality categories. Peak *R*_*t*_ and nadir *R*_*t*_ were defined using (methods). Adjusted hazard ratios and 95% confidence interval of the estimate for risk of COVID-19 were obtained from fully adjusted Cox models.

## Discussion

In this large survey among UK and US participants prospectively assessing risk and severity of COVID-19 infection, we found that a dietary patterns characterized by healthy plant foods was associated with lower risk and severity of COVID-19. We observed a risk gradient of poor diet quality and increased socioeconomic deprivation that departed from the additivity of the risks attributable to each factor separately, suggesting that the beneficial association of diet with COVID-19 may be particularly evident among individuals with higher socioeconomic deprivation.

Our findings are aligned with preliminary evidence showing that improving nutrition could help reduce the burden of infectious diseases.^12,14,24^ Early studies have shown that the administration of arachidonic or linoleic acid partially suppresses SARS-CoV-1 and coronavirus 229E viral replication,^25^ and that specific nutrients or dietary supplements associate with modest reductions in COVID-19 risk.^26^ Results from this observational study could expand previous single nutrient observations and highlight the beneficial association of healthy dietary patterns, which was most pronounced for risk of severe COVID-19. Our findings also concur with a comparative risk assessment study suggesting that a 10% reduction in the prevalence of diet-related conditions such as obesity and type 2 diabetes would have prevented ∼11% of the COVID-19 hospitalizations that have occurred among US adults since November 2020.^27^

The association of healthy diet with lower COVID-19 risk appears particularly evident among individuals living in areas of higher socioeconomic deprivation. Our models estimate that nearly a third of COVID-19 cases would have been prevented if one of two exposures (diet and deprivation) were not present. Although these estimated attributable risks should be interpreted in the context of the population-specific prevalence, and are likely to change over time with the prevailing SARS-CoV-2 infection rate, our observations are consistent with data from ecological studies showing that people living in regions with greater social inequalities are likely to have higher rates of COVID-19 incidence and deaths.^28^ By generating a granular deprivation index based on zip code information our study adds to previous country-level ecological studies. In addition, recent studies on the impact of socioeconomic status on COVID-19 have shown that community-level deprivation indices are strongly associated with COVID-19 risk and mortality.^29,30^ However, it is still possible that differences in deprivation exists within communities. Further studies including information about household characteristics, built environment, or access to healthy foods are needed to expand these initial associations.

Our study adds to knowledge by formally investigating how diet quality, in the context of distal social determinants of health, associates with risk and severity of COVID-19. While our study supports the beneficial association of diet quality with COVID-19 risk and severity, particularly among individuals with higher deprivation, we cannot completely rule out the potential for residual confounding. Individuals who eat healthier diets are likely to share other features that might be associated with lower risk of infection such as the adoption of other risk mitigation behaviors, better household conditions and hygiene, or access to care. However, it is reassuring that our findings were consistent despite controlling for additional surrogate markers of SARS-CoV-2 infection such as mask wearing or community transmission rate, two of the most relevant factors associated with virus transmission and COVID-19 risk.^31^ These findings suggest that efforts to address disparities in COVID-19 risk and severity should consider specific attention to access to healthy foods as a social determinants of health.

We acknowledge several limitations. First, as an observational study, we are unable to confirm a direct causal association between diet and COVID-risk or infer specific mechanisms. Second, our study population was not a random sampling of the population. Although this limitation is inherent to any study requiring voluntary provision of information, we recognize our participants are mainly white participants and less likely to live in low deprived areas and are less ethnically diverse than the general population.^19^ Thus, generalizability of our finding even to the wider British and American population is uncertain. Third, our results could be biased due to the time lapse between the dietary recalls, administered a few months after the relevant period of exposure (pre-pandemic). However, our sensitivity analyses in which we censored cases that had occurred before the administration of the diet survey showed consistent results. Fourth, the self-reported nature of the diet questionnaire is prone to measurement error and bias, and the use of a short food frequency survey could have further reduced the resolution of dietary data collected. More accurate dietary intake assessment methods such as the use of dietary intake biomarkers would be valuable in future studies,^32^ but also difficult to implement in large-scale and time-sensitive investigations. Fifth, we defined risk of severe COVID-19 according to reports of hospitalization with oxygen support, which may not have captured more severe or fatal cases.

## Conclusions

In conclusion, our data provide evidence that a healthy diet was associated with lower risk of COVID-19 and severe COVID-19 even after accounting for other healthy behaviors, social determinants of health, and virus transmission measures. The joint association of diet quality with socioeconomic deprivation was greater than the addition of the risks associated with each individual factor, suggesting that diet quality may play a direct influence in COVID-19 susceptibility and progression. Our findings suggest that public health interventions to improve nutrition and poor metabolic health and address social determinants of health may be important for reducing the burden of the pandemic.

## Data Availability

Data availability statement
Zoe Platform data used in this study is available to researchers through UK Health Data Research using the following link:
https://web.www.healthdatagateway.org/dataset/fddcb382-3051-4394-8436-b92295f14259. The diet quality data used for this study are held by the department of Twin Research at Kings College London. The data can be released to bona fide researchers using our normal procedures overseen by the Wellcome Trust and its guidelines as part of our core funding. We receive around 100 requests per year for our datasets and have a meeting three times a month with independent members to assess proposals. Application is via https://twinsuk.ac.uk/resources-for-researchers/access-our-data/. This means that the data needs to be anonymized and conform to GDPR standards.

## Acknowledgments

We express our sincere thanks to all of the participants who entered data into the app, including study volunteers enrolled in cohorts within the Coronavirus Pandemic Epidemiology (COPE) consortium. We thank the staff of Zoe Global, the Department of Twin Research at King’s College London and the Clinical and Translational Epidemiology Unit at Massachusetts General Hospital for tireless work in contributing to the running of the study and data collection. This work was conducted using the Short Form FFQ tool developed by Cleghorn as reported in https://doi.org/10.1017/S1368980016001099 and listed in the Nutritools (www.nutritools.org) library.

## Contributions

JM, ADJ, LHN, ERL, TDS, SEB, and ATC conceived the study design. JM, ADJ, ERL, MSG, JC, BM, SS contributed to the statistical analysis. All authors were involved in acquisition, analysis, or interpretation of data. JM, LHN, and DAD wrote the first draft of the manuscript. DAD, WCW, SO, CJS, JW, PWF, TDS, SEB, ATC obtained funding. JM, ADJ, LHN provided administrative, technical, or material support. TDS, SEB, ATC jointly supervised this work. All authors contributed to the critical revision of the manuscript for important intellectual content and approved the final version of the manuscript. The corresponding authors attest that all listed authors meet authorship criteria and that no others meeting the criteria have been omitted.

## Data availability statement

Zoe Platform data used in this study is available to researchers through UK Health Data Research using the following link:

https://web.www.healthdatagateway.org/dataset/fddcb382-3051-4394-8436-b92295f14259. The diet quality data used for this study are held by the department of Twin Research at Kings’ College London. The data can be released to bona fide researchers using our normal procedures overseen by the Wellcome Trust and its guidelines as part of our core funding. We receive around 100 requests per year for our datasets and have a meeting three times a month with independent members to assess proposals. Application is via https://twinsuk.ac.uk/resources-for-researchers/access-our-data/. This means that the data needs to be anonymized and conform to GDPR standards.

