## Supplement for "Diet quality and risk and severity of COVID-19: a prospective cohort study"

**Table of Contents:**

**Supplementary Methods**

Dietary intake assessment	3

Outcome ascertainment	3

Generation of socioeconomic measures	3

Covariate classification	4

Interactions between diet quality and deprivation on COVID-19 risk	4

*R_t_* data extraction and definitions	4

Mask wearing	4

References	5

**Supplementary Tables**

Supplementary table 1: Grouping and components of the hPDI	6

Supplementary table 2: Criteria for scoring each component of the hPDI	7

Supplementary table 3: Grouping and components of the DQS	8

Supplementary table 4: Criteria for scoring each component of the DQS	9

Supplementary table 5: Adjusted hazard ratios of COVID-19 risk and severity for diet quality in the COVID Symptom Study	10

Supplementary table 6: Association between diet quality and COVID risk - censored to cases that occurred after completing the diet questionnaires	12

Supplementary table 7: Attributing associations to additive interaction between diet quality and socioeconomic deprivation on risk of COVID-19 infection	13

Supplementary table 8: Association between diet quality and risk of COVID-19 infection after accounting for mask wearing	14

**Supplementary Figures**

Supplementary figure 1: Flow diagram	15

Supplementary figure 2: Distribution of the hPDI score	16

Supplementary figure 3: Dose-response associations between diet quality and risk of COVID-19 infection.	17

Supplementary figure 4: Absolute excess rate of COVID-19 per 10,000 person-months according to socioeconomic deprivation and diet quality	18

**Supplementary Methods**

**Dietary intake assessment**

Habitual dietary intake information was collected through an amended version of the Leeds Short Form Food Frequency Questionnaire (LSF-FFQ). ^1^ In brief, the LSF-FFQ includes 20 food items with reference to fruit, vegetables, fibre-rich foods, high fat and high-sugar foods, meat, meat products and fish. Seven additional food items were added to capture broader dietary intake information including refined carbohydrates (e.g. white rice, white pasta and white bread), eggs, fast food and live probiotic or fermented foods (e.g.  live yogurt, kefir and kimchi). Participants were asked how often on average they had consumed one portion of each food in a typical week. The responses had eight frequency categories ranging from “rarely or never” to “five or more times per day. Further detail on the development, dissemination and procedures of the diet and lifestyle survey to UK and US participants is described elsewhere.^2^

**Outcome ascertainment**

Predicted COVID-19 definition: We used a symptom-based classifier developed by our group to predict COVID-19.^3^ To build the prediction model, UK participants were randomly divided into a training set and a test set (ratio: 80:20). Based on the training set, a logistic model generated to predict symptomatic COVID-19 was: Log odds (Predicted COVID-19) = -1.32 - (0.01 x age) + (0.44 x male sex) + (1.75 x loss of smell or taste) + (0.31 x severe or significant persistent cough) + (0.49 x severe fatigue) + (0.39 x skipped meals). The prediction model achieved a sensitivity of 0.65 (95% CI 0.62-0.67) and specificity of 0.78 (95% CI 0.76-0.80) in the test set. In additional validation in the U.S. participants, the prediction model achieved a sensitivity of 0.66 (95% CI 0.62-0.69) and specificity of 0.83 (95% CI 0.82-0.85).

Severe COVID: To ascertain severe COVID-19, we used responses to the question “*What treatment did you receive while in the hospital / What treatment are you receiving right now?*” Participants had the option to respond a) None, b) Oxygen and fluids breathing support administered through an oxygen mask, no pressure applied, c) Non-invasive ventilation breathing support administered through an oxygen mask, which pushes oxygen into your lungs, d) Invasive ventilation breathing support administered through an inserted tube. People are usually asleep for this procedure, e) Other. COVID-19 severity was ascertained based on a report of the need for a hospital visit which required 1) non-invasive breathing support, 2) invasive breathing support, and 3) administration of antibiotics combined with oxygen support.

**Generation of socioeconomic measures**

Based on zip code or post code of residence, participants were assigned to community-level socioeconomic measures and population density.

For participants in the US, socioeconomic measures were generated using aggregated census data for up to 25 characteristics that have been used consistently to approximate neighborhood-level environments^4^. Principal component analysis was used for census data reduction and seven variables were retained for the generation of the index. Loads were obtained from the principal component analysis. The deprivation index was then standardized to have a mean of 0 and a standard deviation of 1.

For UK participants we retrieved the education and income measures for the indices of multiple deprivation calculated by the Office of National Statistics from 2019 data^5^ aggregated to the 2011 Lower Super Output Areas. Population density was calculated from Census data for all Zip Code Tabulation Areas in the U.S and from data calculated by the Office of National Statistics, from 2019 data^6^, again aggregated to 2011 Lower Super Output Areas.

**Covariate classification**

Covariates were selected *a priori* based on putative confounders and risk factors for COVID-19 and included sex (male, female), race/ethnicity (White, Black, Asian, Other), index of multiple deprivation (most deprived <3, intermediate deprived 3 to 7, less deprived >7), population density (<500 individuals/km^2^, 500 to 1,999 individuals/km^2^, 2,000 to 4,999 individuals/km^2^, and ≥ 5,000 individuals/km^2^), healthcare worker status (yes with interaction with COVID-19 patients, yes without interaction with COVID-19 patients, no), presence of comorbidities [diabetes (yes, no), cardiovascular disease (yes, no), lung disease (yes, no), cancer (yes, no), kidney disease (yes, no)], body mass index (<18.5 kg/m2, 18.5 to 24.9 kg/m2, 25.0 to 29.9 kg/m2, and ≥30 kg/m2), smoking status (yes, no), and physical activity (<1 day/week, 1 to 2 days/week, 3 to 4 days/week, ≥5 days/week).

**Interactions between diet quality and deprivation on COVID-19 risk**

We tested for additive interactions by assessing the relative excess risk due to interaction, and further examined the risk proportions attributable to diet quality alone, to deprivation alone, and to their interaction. For these analyses, we considered diet quality and socioeconomic deprivation as continuous variables. We assessed the relative excess risk due to interaction as an index of additive interaction using the following formula (RERI = RR_11_ - RR_10_ - RR_01_ + 1)^7^, and further examined the decomposition of the joint effect, which is the proportion attributable to genetic risk alone, to diet quality alone, and to their interaction (i.e., AP= RERI/ RR_11_).^7^

***R_t_* data extraction and definitions**

We extracted US state-level information on *R_t_* from the COVID Tracking Project,^8^ for the period between March 2020 and January 2021. For the UK we calculated *R_t_* time-series for Scotland, Wales, and each of the NHS regions in England, using previously published methodology^9^. In brief, newly sick users of the app were invited to take Covid-19 tests. These test results was used to produce regional-level incidence estimates, from which we estimated *R_t_* using a probabilistic model. For these analyses, we defined community peak and nadir *R_t_* time-windows as the period between one week before and two weeks after *R_t_* was all-time high/low. Using censored time-windows, we tested the association between diet quality and COVID-19 risk after adjusting for the same confounders as included in model 3.

**Mask wearing**

Beginning in June 2020 and until September 2020, study participants were also asked to report on each entry whether they had worn a face mask when outside the house in the last week and categorized responses into never, sometimes, most of the time, or always. For mask wearing analyses, which included the same covariates as included in model 3, we collapsed responders into two categories: participants who wore masks ‘none of the time or sometimes’, and those who reported wearing masks ‘most of time/ always’ at least once.

6 [Small Area Population Estimates: Summary of methodology review and research update - Office for National Statistics](https://secure-web.cisco.com/1sWIjvn8gpUlUxCk8sZHBCfEaawgaxqxptg7uWJ6ausLzAAkXmwITmjeXzJTnWEK7lsXaXEkVvNDIJQtByEks2WJp8ISE82rsEV9l2oT1vvzeSNeq_zMmm5BbcB1IFCRY1PgbiAC8-SXXubeg6m1V21FsK0iWnOf--6E7qLTWJq9geHfMRI_LBZgDdM7KADkSSKRtRdvp-sIaL29WCc-38_FZb-zGNRaGlJnfjLJ5HbVfCB2hYTHFdqtpZ43fkIEb7wmkPHLlJ06s7QaO1a-x0g/https%3A%2F%2Fwww.ons.gov.uk%2Fpeoplepopulationandcommunity%2Fpopulationandmigration%2Fpopulationestimates%2Fmethodologies%2Fsmallareapopulationestimatessummaryofmethodologyreviewandresearchupdate) <https://www.ons.gov.uk/peoplepopulationandcommunity/populationandmigration/populationestimates/methodologies/smallareapopulationestimatessummaryofmethodologyreviewandresearchupdate> [Date accessed 2021 Feb 02]

7 VanderWeele TJ, Tchetgen Tchetgen EJ. Attributing effects to interactions. Epidemiology 2014;25:711–22.

8 The Covid Tracking Project. <https://covidtracking.com> [Date accessed 2020 Dec 06]

9 Varsavsky T, Graham MS, Canas LS, et al. Detecting COVID-19 infection hotspots in England using large-scale self-reported data from a mobile application: a prospective, observational study. Lancet Public Heal 2021;6:e21–e29.

**Supplementary table 1: Grouping and components of the hPDI**

| **hPDI component** | **FFQ items** |
| --- | --- |
| Wholegrains | Fibre-rich breakfast cereal, like Weetabix, Fruit ‘n Fibre, Porridge, Muesli; Wholemeal bread or chapattis |
| Fruits | Fruit (tinned / fresh) |
| Vegetables | Salad (not garnish added to sandwiches); Vegetables (tinned / frozen / fresh but not potatoes) |
| Nuts | N/A |
| Legumes | Beans or pulses like baked beans, chick peas, dahl |
| Vegetable oils | N/A |
| Tea and Coffee | N/A |
| Fruit Juice | Fruit juice (not cordial or squash) |
| Refined Grains | Crisps / savoury snacks; pasta; Refined breakfast cereals (e.g. rice krispies, cornflakes, coco pops); rice; white bread |
| Potatoes | Chips / fried potatoes |
| Sugar Sweetened Beverages | Nonalcoholic fizzy drinks/pop  (not sugar free or diet) |
| Sweets and desserts | Sweet biscuits, cakes, chocolate, sweets |
| Animal fats | N/A |
| Dairy | Cheese / yoghurt; Ice cream / cream; live probiotic or fermented food products (e.g. yoghurt, kefir, kimchi) |
| Egg | Eggs - as boiled, fried, scrambled, etc |
| Fish and seafood | White fish in batter or breadcrumbs – like ‘fish ‘n chips’; White fish not in batter or breadcrumbs; Oily fish – like herrings, sardines, salmon, trout, mackerel, fresh tuna (not tinned tuna) |
| Meat | Beef, Lamb, Pork, Ham - steaks, roasts, joints, mince or chops; Chicken or Turkey – steaks, roasts, joints, mince or portions (not in batter or breadcrumbs); Sausages, bacon, corned beef, meat pies/pasties, burgers; Chicken/turkey nuggets/twizzlers, turkey burgers, chicken pies, or in batter or breadcrumbs |
| Miscellaneous | Fast food |

**Table legend:** FFQ items constituting the 18 food groups originally used to generate the healthy plant-based diet index in Satija et al. JACC 2017. Four out of the 18 food groups originally considered were not included for the calculation of the healthy plant-based diet index in this study as they were not available (N/A). FFQ = food frequency questionnaire; hPDI = healthful plant-based diet index

**Supplementary table 2: Criteria for scoring each component of the hPDI**

| **Component** | **Criteria for min score of 1** | **Criteria for max score of 5** |
| --- | --- | --- |
| Whole grain | Lowest quintile of intake | Highest quintile of intake |
| Fruits | Lowest quintile of intake | Highest quintile of intake |
| Vegetables | Lowest quintile of intake | Highest quintile of intake |
| Nuts | N/A | N/A |
| Legumes | Lowest quintile of intake |  |
| Vegetable oils | N/A | N/A |
| Tea and coffee | N/A | N/A |
| Fruit juices | Highest quintile of intake | Lowest quintile of intake |
| Refined grains | Highest quintile of intake | Lowest quintile of intake |
| Potatoes | Highest quintile of intake | Lowest quintile of intake |
| Sugar sweetened beverages | Highest quintile of intake | Lowest quintile of intake |
| Sweets and desserts | Highest quintile of intake | Lowest quintile of intake |
| Animal fat | N/A | N/A |
| Dairy | Highest quintile of intake | Lowest quintile of intake |
| Egg | Highest quintile of intake | Lowest quintile of intake |
| Fish or seafood | Highest quintile of intake | Lowest quintile of intake |
| Meat | Highest quintile of intake | Lowest quintile of intake |
| Miscellaneous | Highest quintile of intake | Lowest quintile of intake |

**Table legend:** Criteria for scoring the 18 food groups originally used to generate the healthy plant-based diet index in Satija et al. JACC 2017. Food groups were ranked into quintiles, and given positive (healthy plant food groups) or reverse scores ( less healthy plant food groups and animal food groups). With positive scores, participants above the highest quintile of a food group received a score of 5, following on through to participants below the lowest quintile who received a score of 1. With reverse scores, this pattern of scoring was inverted. All component scores were summed to obtain a total score ranging from 0 (lowest diet quality) to 70 (highest diet quality) points.

**Supplementary table 3: Grouping and components of the DQS**

| **DQS component** | **FFQ items** |
| --- | --- |
| Fruits | Fruit (tinned / fresh) |
| Vegetables | Salad (not garnish added to sandwiches) Vegetables (tinned / frozen / fresh but not potatoes) |
| Oily fish | Oily fish – like herrings, sardines, salmon, trout, mackerel, fresh tuna (not tinned tuna) |
| Total fat | Fruit (tinned / fresh) Fruit juice (not cordial or squash) Salad (not garnish added to sandwiches) Vegetables (tinned / frozen / fresh but not potatoes) Chips / fried potatoes Beans or pulses like baked beans, chick peas, dahl Fiber-rich breakfast cereal, like Weetabix, Fruit ‘n Fiber, Porridge, Muesli Whole-meal bread or chapattis; Cheese / yoghurt; Crisps / savory snacks Sweet biscuits, cakes, chocolate, sweets Ice cream / cream Nonalcoholic fizzy drinks/pop (not sugar free or diet) Beef, Lamb, Pork, Ham - steaks, roasts, joints, mince or chops Chicken or Turkey – steaks, roasts, joints, mince or portions (not in batter or breadcrumbs) Processed meats/ meat products Sausages, bacon, corned beef, meat pies/pasties, burgers Chicken/turkey nuggets/twizzles, turkey burgers, chicken pies, or in batter or breadcrumbs White fish in batter or breadcrumbs – like ‘fish ‘n chips’ White fish not in batter or breadcrumbs |
| Non-milk extrinsic sugars | Fruit (tinned / fresh) Fruit juice (not cordial or squash) Salad (not garnish added to sandwiches) Vegetables (tinned / frozen / fresh but not potatoes) Chips / fried potatoes Beans or pulses like baked beans, chick peas, dahl Fiber-rich breakfast cereal, like Weetabix, Fruit ‘n Fiber, Porridge, Muesli Whole-meal bread or chapattis; Cheese / yoghurt; Crisps / savory snacks Sweet biscuits, cakes, chocolate, sweets Ice cream / cream Nonalcoholic fizzy drinks/pop (not sugar free or diet) Beef, Lamb, Pork, Ham - steaks, roasts, joints, mince or chops Chicken or Turkey – steaks, roasts, joints, mince or portions (not in batter or breadcrumbs) Processed meats/ meat products Sausages, bacon, corned beef, meat pies/pasties, burgers Chicken/turkey nuggets/twizzles, turkey burgers, chicken pies, or in batter or breadcrumbs White fish in batter or breadcrumbs – like ‘fish ‘n chips’ White fish not in batter or breadcrumbs |

**Table legend:** FFQ items constituting the 5 food components originally used to generate the DQS score from Cleghorn et al., listed in the Nutritools library (nutritools.org). FFQ = food frequency questionnaire; DQS = diet quality score.

**Supplementary table 4: Criteria for scoring each component of the DQS**

| **DQS Component** | **Criteria for score of 1** | **Criteria for score of 2** | **Criteria for score of 3** |
| --- | --- | --- | --- |
| Fruit | ≤ 2 servings/week | >2 servings/week and <2 servings/d | ≥2 servings/d |
| Vegetables | ≤ 1 servings/d | 1-3 servings/d | ≥ 3 servings/d |
| Oily Fish | No intake | 0-200g/week | 200g/week |
| Total Fat | ≥1.5 x UK recommendations  ( ≥127.5g/d) | 1-1.5 x UK recommendations | ≤ UK recommendations  (≤ 85g/d) |
| Non-Milk-Extrinsic Sugars | ≥1.5 x UK recommendations  ( ≥ 90g/d) | 1-1.5 x UK recommendations | ≤ UK recommendations  (≤ 60g/d) |

**Table legend:** Criteria for scoring the 5 food groups originally used to generate the diet quality score from Cleghorn et al., listed in the Nutritools library (nutritools.org). Each component was scored from 1 (unhealthiest) to 3 (healthiest) points, with intermediate values scored proportionally. All component scores were summed to obtain a total score ranging from 5 (lowest diet quality) to 15 points(highest diet quality) points.

**Supplementary table 5: Adjusted hazard ratios of COVID-19 risk and severity for diet quality in the COVID Symptom Study**

|  | **Low DQS** | **Intermediate DQS** | **High DQS** | ***P* for trend** |
| --- | --- | --- | --- | --- |
| Diet quality score, median (IQR) | 9 (8-10) | 11 (11-11) | 13 (12 -13) |  |
| **COVID-19 risk** |  |  |  |  |
| No. of events/person-months | 13,996 / 1,467,205 | 12,641 / 1,701,799 | 5,178 / 717,270 | — |
| Incidence rate (10,000 person-months; 95% CI) | 95.4 (93.8-97.0) | 74.3 (73.0-75.6) | 72.2 (70.3-74.2) | — |
| Age-adjusted model | 1.00 (Ref) | 0.90 (0.87-0.92) | 0.92 (0.89-0.95) | <0.001 |
| Multivariable model 2 | 1.00 (Ref) | 0.90 (0.88-0.92) | 0.92 (0.89-0.95) | 0.019 |
| Multivariable model 3 | 1.00 (Ref) | 0.95 (0.93-0.98) | 1.00 (0.97-1.03) | 0.216 |
| **COVID-19 risk (positive test)** |  |  |  |  |
| No. of events/person-months | 2,341 / 1,515,004 | 2,309 / 1,746,982 | 952 / 736,535 | — |
| Incidence rate (10,000 person-months; 95% CI) | 15.5 (14.8-16.1) | 13.2 (12.7-13.8) | 12.9 (12.1-13.7) | — |
| Age-adjusted model^$^ | 1.00 (Ref) | 0.94 (0.91-0.98) | 0.92 (0.87-0.96) | <0.001 |
| Multivariable model 2^$^ | 1.00 (Ref) | 0.95 (0.91-0.99) | 0.93 (0.89-0.98) | 0.006 |
| Multivariable model 3^$^ | 1.00 (Ref) | 0.96 (0.93-1.00) | 0.95 (0.91-1.00) | 0.047 |
| **COVID-19 severity** |  |  |  |  |
| No. of events/person-months | 313 / 1,518,980 | 317 / 1,750,786 | 110 / 738,495 | — |
| Incidence rate (10,000 person-months; 95% CI) | 2.1 (1.8-2.3) | 1.8 (1.6-2.0) | 1.5 (1.2-1.8) | — |
| Age-adjusted model | 1.00 (Ref) | 0.82 (0.70-0.96) | 0.67 (0.54-0.84) | <0.001 |
| Multivariable model 2 | 1.00 (Ref) | 0.82 (0.70-0.97) | 0.68 (0.54-0.85) | <0.001 |
| Multivariable model 3 | 1.00 (Ref) | 0.93 (0.79-1.09) | 0.83 (0.66-1.04) | 0.141 |

**Table legend:** Hazards ratios and 95% CI for COVID-19 risk and severity. Sensitivity analysis using the DQS to quantify diet quality. Cox proportional hazards models were stratified by calendar date at study entry, country of origin, and 10-year age group (Age-adjusted model). Multivariable model 2 was further adjusted for sex (male, female), race/ethnicity (White, Black, Asian, Other), index of multiple deprivation (most deprived <3, intermediate deprived 3 to 7, less deprived >7), population density (<500 individuals/km^2^, 500 to 1,999 individuals/km^2^, 2,000 to 4,999 individuals/km^2^, and ≥ 5,000 individuals/km^2^), and healthcare worker status (yes with interaction with COVID-19 patients, yes without interaction with COVID-19 patients, no). Model 3 was further adjusted for presence of comorbidities [diabetes (yes, no), cardiovascular disease (yes, no), lung disease (yes, no), cancer (yes, no), kidney disease (yes, no)], body mass index (<18.5 kg/m2, 18.5 to 24.9 kg/m2, 25.0 to 29.9 kg/m2, and ≥30 kg/m2), smoking status (yes, no), and physical activity (<1 day/week, 1 to 2 days/week, 3 to 4 days/week, ≥5 days/week).

^$^ Inverse probability-weighted analyses were conducted to account for predictors of obtaining RT-PCR testing (presence of COVID-19-related symptoms, interaction with a COVID-19 case, healthcare worker, age group, and race). inverse probability-weighted Cox proportional hazards models were stratified by 10-year age group and date with additional adjustment for the covariates used in previous models.

**Supplementary table 6: Association between diet quality and COVID risk - censored to cases that occurred after completing the diet questionnaires**

|  | **Low hPDI** | **Intermediate hPDI** | **High hPDI** | ***P* for trend** |
| --- | --- | --- | --- | --- |
| **COVID-19 risk** |  |  |  |  |
| Incidence rate (10,000 person-months; 95% CI) | 116.2 (110.9-120.3) | 84.4 (82.0-86.7) | 74.1 (71.5-78.6) | ~~—~~ |
| Age-adjusted model | 1.00 (Ref) | 0.82 (0.78-0.86) | 0.79 (0.75-0.83) | <0.001 |
| Multivariable model 2 | 1.00 (Ref) | 0.83 (0.79-0.87) | 0.79 (0.75-0.84) | <0.001 |
| Multivariable model 3 | 1.00 (Ref) | 0.87 (0.83-0.92) | 0.88 (0.83-0.93) | <0.001 |
| **COVID-19 risk (positive test)** |  |  |  |  |
| Incidence rate (10,000 person-months; 95% CI) | 42.1 (40.2-44.3) | 33.5 (32.1-35.0) | 29.1 (27.2-31.0) | ~~—~~ |
| Age-adjusted model^$^ | 1.00 (Ref) | 0.84 (0.77-0.92) | 0.77 (0.70-0.86) | <0.001 |
| Multivariable model 2^$^ | 1.00 (Ref) | 0.85 (0.78-0.93) | 0.79 (0.72-0.88) | <0.001 |
| Multivariable model 3^$^ | 1.00 (Ref) | 0.86 (0.79-0.94) | 0.80 (0.72-0.89) | <0.001 |

**Table legend:** Hazards ratios and 95% CI for COVID-19 risk. Sensitivity analysis censored to cases that occurred after completing the diet questionnaires (September 21^st^, 2020). Cox proportional hazards models were stratified by calendar date at study entry, country of origin, and 10-year age group (Age-adjusted model). Multivariable model 2 was further adjusted for sex (male, female), race/ethnicity (White, Black, Asian, Other), index of multiple deprivation (most deprived <3, intermediate deprived 3 to 7, less deprived >7), population density (<500 individuals/km^2^, 500 to 1,999 individuals/km^2^, 2,000 to 4,999 individuals/km^2^, and ≥ 5,000 individuals/km^2^), and healthcare worker status (yes with interaction with COVID-19 patients, yes without interaction with COVID-19 patients, no). Model 3 was further adjusted for presence of comorbidities [diabetes (yes, no), cardiovascular disease (yes, no), lung disease (yes, no), cancer (yes, no), kidney disease (yes, no)], body mass index (<18.5 kg/m2, 18.5 to 24.9 kg/m2, 25.0 to 29.9 kg/m2, and ≥30 kg/m2), smoking status (yes, no), and physical activity (<1 day/week, 1 to 2 days/week, 3 to 4 days/week, ≥5 days/week).

^$^ Inverse probability-weighted analyses were conducted to account for predictors of obtaining RT-PCR testing (presence of COVID-19-related symptoms, interaction with a COVID-19 case, healthcare worker, age group, and race). inverse probability-weighted Cox proportional hazards models were stratified by 10-year age group and date with additional adjustment for the covariates used in previous models.

**Supplementary table 7: Attributing associations to additive interaction between diet quality and socioeconomic deprivation on risk of COVID-19 infection**

|  | **Predicted COVID-19 infection** |
| --- | --- |
| **Main effects** | |
| Diet quality, per 10 units decrease | 1.05 (1.01-1.09) |
| Deprivation index, per category decrease | 1.06 (1.01-1.12) |
| Joint effect | 1.15 (1.09-1.21) |
| **Relative excess risk due to interaction** | |
| Relative excess risk due to interaction | 0.05 (0.02-0.08) |
| *P* | 0.005 |
| **Attributable proportion, %** | |
| Diet quality | 31.9 (18.2-45.6) |
| Deprivation index | 38.4 (26.5-50.3) |
| Additive interaction | 29.7 (2.1-57.3) |

**Table Legend:** Multivariable-adjusted risk of predicted COVID-19 infection estimated from fully adjusted Cox models. The relative excess risk due to interaction was calculated using the following formula (RERI_RR_ = RR_11_ - RR_10_ - RR_01_ + 1). The decomposition of the joint effect, which is the proportions attributable to diet quality alone, to deprivation index alone, and to their interaction, was calculated using the following formula (i.e., AP= RERI / RR_11_).

**Supplementary table 8: Association between diet quality and risk of COVID-19 infection after accounting for mask wearing**

|  | **Low hPDI** | **Intermediate hPDI** | **High hPDI** | ***P* for trend** |
| --- | --- | --- | --- | --- |
| **COVID-19 risk** |  |  |  |  |
| No. of events/person-months | 2,574 / 222,426 | 4,669 / 555,918 | 2,092 / 283,975 | — |
| Incidence rate (10,000 person-months; 95% CI) | 114.6 (110.2-119.0) | 84.0 (81.6-86.4) | 73.7 (70.6-76.9) | ~~—~~ |
| Multivariable Model 3 + Mask wearing | 1.00 (Ref) | 0.88 (0.83 to 0.92) | 0.88 (0.83-0.94) | <0.001 |
| **COVID-19 risk (positive test)** |  |  |  |  |
| No. of events/person-months | 989 / 233,564 | 1,907 / 576,267 | 874 / 293,760 |  |
| Incidence rate (10,000 person-months; 95% CI) | 42.3 (39.8-45.1) | 33.1 (31.6-34.6) | 29.8 (27.8-31.8) |  |
| Multivariable Model 3 + Mask wearing^$^ | 1.00 (Ref) | 0.86 (0.79-0.94) | 0.80 (0.72-0.89) | <0.001 |

**Table legend:** Hazards ratios and 95% CI for COVID-19 risk after accounting for mask wearing. These analyses were left censored to September 21^st^ 2020. Mask wearing analyses included 524,825 participants. For confirmed COVID-19 analyses, inverse probability-weighted analyses were conducted to account for predictors of obtaining RT-PCR testing^$^.

**Supplementary figure 1: Flow diagram**

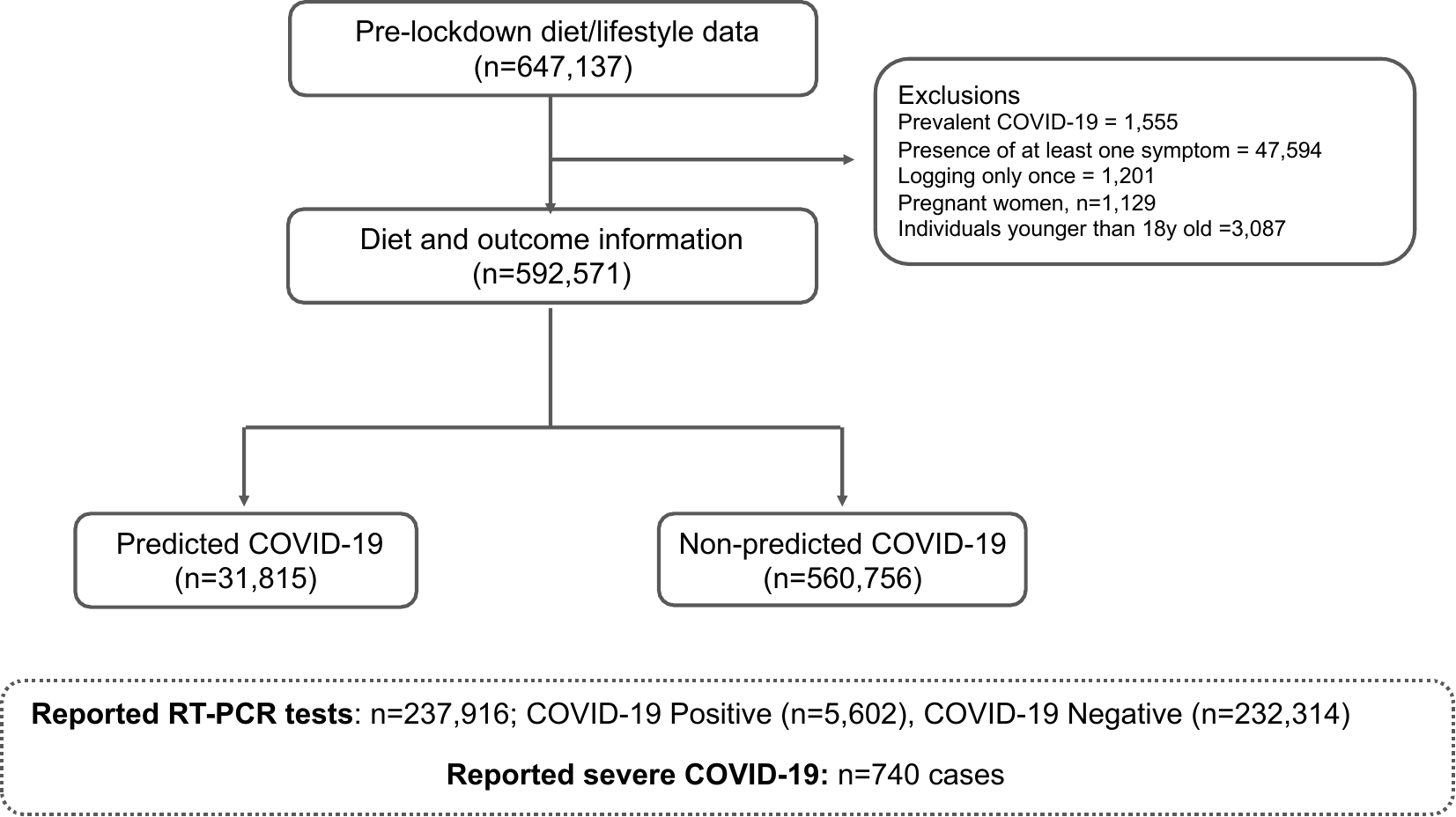

**Figure legend:** Identification of participants with diet and lifestyle data at baseline who met the eligibility criteria for this study. Number of cases and controls identified until the end of follow-up (December 2^nd^, 2020)

**Supplementary figure 2: Distribution of the hPDI score**

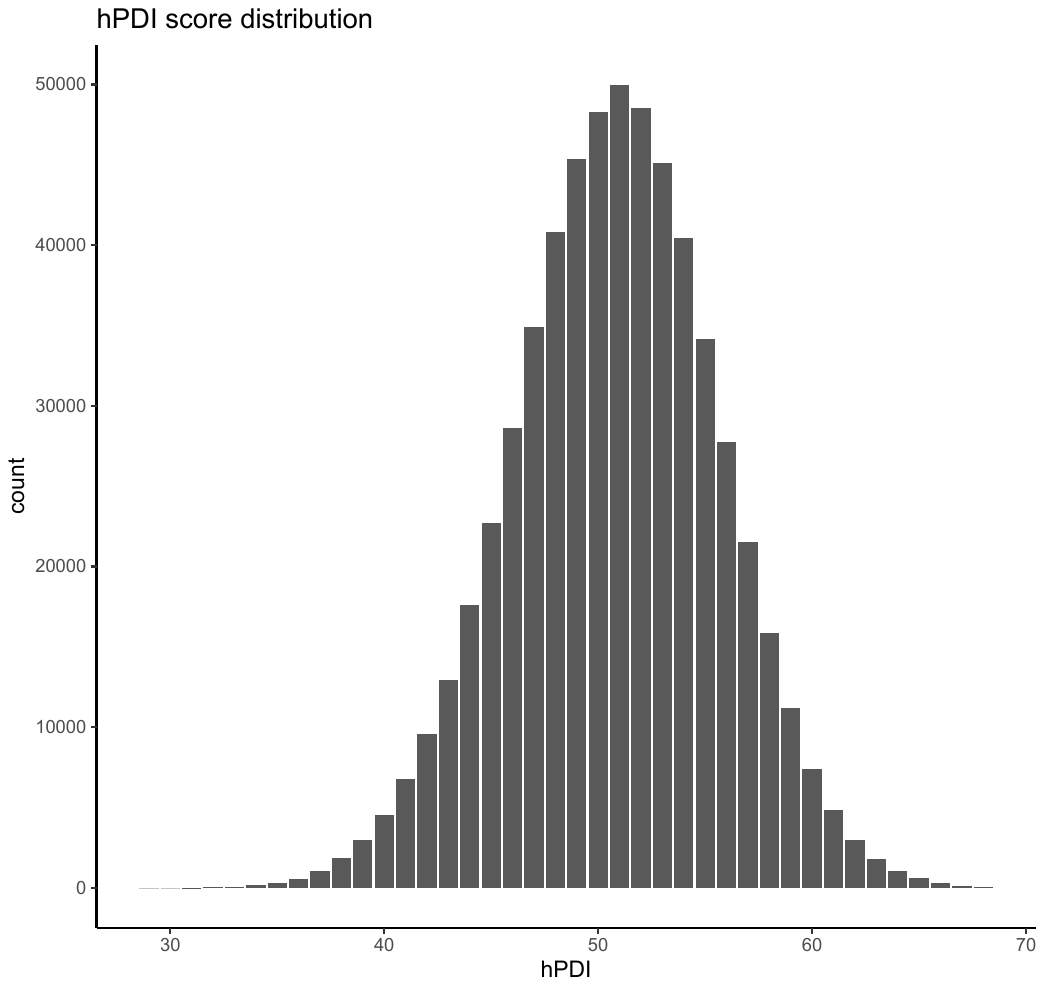

**Figure legend:** Distribution of the healthy plant-based diet index.

**Supplementary figure 3: Dose-response associations between diet quality and risk of COVID-19 infection.**

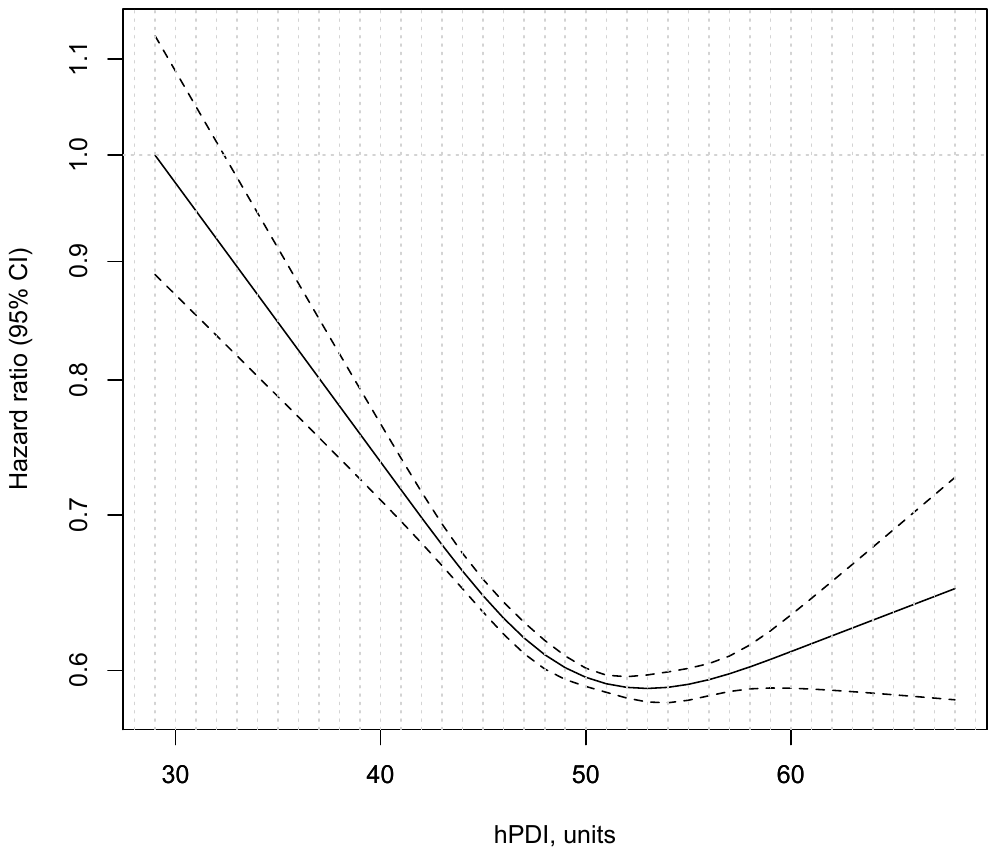

**Figure legend:** Dose-response associations between diet quality and risk of COVID-19 infection were calculated using restricted cubic splines with four knots (at the 2.5th, 25th, 75th, and 97.5th percentiles; methods). Cox models were adjusted for all confounders previously included in model 3. P for non-linearity <0.001.

**Supplementary figure 4: Absolute excess rate of COVID-19 per 10,000 person-months according to socioeconomic deprivation and diet quality**

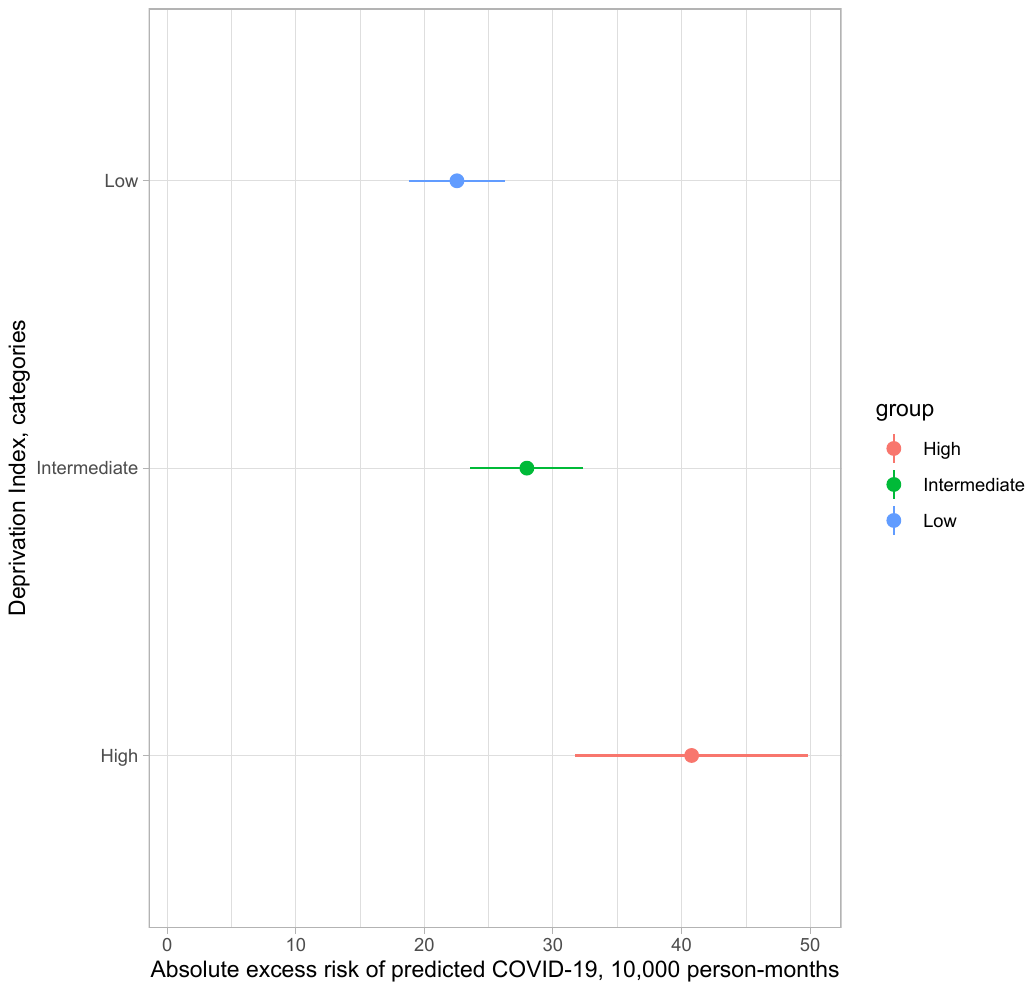

**Figure legend:** Absolute excess risk of COVID-19 per 10,000 person-months for lowest vs highest quartile of the diet score according to socioeconomic deprivation. Absolute excess risk was calculated based on the incidence rate per 1,000 person-months in each diet quality score and socioeconomic deprivation category using the “epiR” package in R.
